# Chloroquine, but not hydroxychlorquine, prolongs the QT interval in a primary care population

**DOI:** 10.1101/2020.06.19.20135475

**Authors:** Jonas L. Isaksen, Anders G. Holst, Adrian Pietersen, Jonas B. Nielsen, Claus Graff, Jørgen K. Kanters

**Author notes:** Address for correspondence: Jørgen K. Kanters, Department of Biomedical Sciences 10.5, Nørre Allé 14, 2200 Copenhagen N, Denmark.

## Abstract

**Background:** Chloroquine (CQ) and Hydroxychloroquine (HCQ) have recently been suggested as treatment for the current Corona Virus Disease 2019 (COVID-19) pandemic. However, despite their long-term use and only few case reports on adverse effects, CQ and HCQ are listed as a known risk of the lethal ventricular arrhythmia Torsade de Pointes and their cardiac safety profile is being questioned. Thus, we aimed to investigate the electrocardiographic and mortality effects of CQ and HCQ in a primary care population.

**Methods:** We used Danish health care registers and electrocardiograms (ECGs) from primary care to define three studies. 1) A paired study of subjects with ECGs before and during use of CQ/HCQ, 2) a matched ECG study of subjects taking CQ/HCQ compared to controls, and 3) a mortality study on people taking HCQ matched to control. In both matched studies, we adjusted for connective tissue diseases, use of QT-prolonging drugs, and cardiac disease. We used the QTc interval as the marker for electrocardiographic safety. In the mortality study, cases were followed from first claimed prescription until 300 days after estimated completion of the last prescription. 95% confidence intervals follow estimates in parenthesis.

**Results:** Use of CQ was associated with a 5.5 (0.7;10) ms increase in QTc in the paired study (n=10). In the matched study (n=28, controls=280), QTc was insignificantly increased in subjects taking CQ by 4.7 (−3.4;13) ms. With a ΔQTc of 1.0 (−5.6;7.5), use of HCQ was not associated with an increased QTc in the paired study (n=32). In the matched study (n=172, controls=1,720), QTc also was not different between groups (p=0.5). In the mortality study (n=3,368), use of HCQ was associated with a hazard ratio of 0.67 (0.43;1.05).

**Conclusions:** In subjects free of COVID-19, we found a small increase in QTc associated with use of chloroquine, but not hydroxychloroquine. We found no increased mortality associated with use of hydroxychloroquine.

## Introduction

The Corona Virus Disease 2019 (COVID-19) pandemic has led to investigation and use of chloroquine (CQ) and hydroxychloroquine (HCQ) in the treatment of COVID-19. CQ and HCQ are 4-aminoquinilines approved for medical use in 1949 and 1955, respectively, and both drugs interfere with important physiological mechanisms that may improve the clinical course of patients with COVID-19. CQ and HCQ are weak bases and accumulate in lysosomes increasing pH which interferes with virus entry and fusion(1). CQ and HCQ also inhibit viral gene expression and post translational modification necessary for viral replication(1). Furthermore, CQ and HCQ have direct anti-inflammatory actions by inhibition of B- and T-cell receptors and especially by decreasing TNF-α and cytokine production (interleukin-1 and interleukin-6)(2). The drawback is that CQ and HCQ have known cardiotoxic effects(3) including vasodilatation, hypotension, hypokalemia, negative inotropy, and arrhythmias. The risk of arrhythmias may partly be mediated through the other cardiotoxic effect and partly due to ion channel blockade. Both CQ and HCQ are known hERG-blockers, but also block a variety of other ion channels including sodium, calcium, I_K1_ potassium and pacemaker funny channels(4). Since CQ and HCQ are old drugs, the approvals were granted long before thorough QT studies were required and only limited ECG safety data exists. Despite the known hERG block, there is a paucity of documented Torsades de Pointes ventricular tachycardia (TdP) during treatment,(5) which may be attributed to the concomitant calcium block protecting against TdP.(6) Only three cases of TdP have been published(7-9), all in patients with concomitant morbidities as cirrhosis, heart failure, or preexisting QT-prolongation.

Only limited electrocardiographic information is available of the effect of CQ/HCQ. A study of 14 healthy subjects receiving 1.5 g of CQ (normal dose in COVID 0.15 g OD(2)) found significant prolongation of the QT interval around 20 ms(10). With HCQ there does not exist any studies with comparable ECG data before and after treatment. Since COVID-19 attacks the heart causing heart failure and even shock, it is unknown whether the arrhythmias seen in COVID-19 patients treated with HCQ/CQ are secondary to the disease or caused by the HCA/CQ treatment. Thus, an investigation of the electrocardiographic and mortality risks associated with CQ/HCQ treatment is needed.

The aim of the present study was to investigate the effects of CQ/HCQ on electrocardiographic parameters in a large primary care population without COVID-19 using Danish health care registers and electronic ECG data.

## Methods

### ECG

The primary population consisted of 449,584 patients with 978,901 ECGs from the Copenhagen General Practitioners’ Laboratory (CGPL). CGPL was the core facility that received referrals from most General Practitioners in the Copenhagen area from 2001-2015 for blood sampling and ECG examination. Data from CGPL was linked with Danish registries for a full patient history and follow-up. The Danish Civil Register contains information on sex, birthday, and emigration/permanent stays abroad. The Danish Register of Patient Records contains information on all hospital contacts (in-patient and out-patient). The Register used International Classification of Diseases, eighth revision (ICD-8) codes until 1993 and ICD-10 codes from 1994. The Danish Register of Medication Statistics holds information on every claimed prescription from Danish pharmacies. The Danish Register of Causes of Death contains information on mortality for all Danish Citizens.

Using the 12SL algorithm version 23 (GE Healthcare, Wauwatosa, WI), we obtained heart rate, QT interval, QRS duration, JT interval, PR interval, and presence of Right Bundle Branch Block (RBBB) or Left Bundle Branch Block (LBBB). The QT interval was corrected with Bazett (QTcB) and Framingham (QTcFrh) formulae. Based on the JT interval, JTc was obtained by linear correction for heart rate (JTc = JT – 119.4108*(RR-1), whereby RR is the R-to-R interval in seconds and JT is measured in ms).

### Studies and populations

From the primary study population, we defined three subsets, each with a CQ and a HCQ population: 1) Paired study, 2) Matched study, and 3) Mortality study. As an additional analysis, we conducted a fourth study on connective tissue diseases with three populations independent of CQ/HCQ prescriptions.

### Paired study

The Paired study is comprised of people with an ECG taken during a period of CQ or HCQ treatment, respectively, and with a prior baseline ECG without HCQ/CQ treatment. Thus, in this study, the subjects served as their own controls.

### Matched study

The Matched study is a comparison of subjects with an ECG taken during use of CQ or HCQ, respectively, compared to a matched reference population with no history of CQ and HCQ usage, respectively. Matching was done 10:1 on sex and age (rounded to integer years), sampled without replacement.

### Mortality study

The Mortality study is a study of subjects with a prescription of HCQ matched 1:1 to sex- and age-matched controls with no prescription. Cases and their matched control were followed from the day of the first prescription until 300 days after predicted exhaustion of the last claimed prescription of the case. Since HCQ replaced CQ as the preferred drug in 1995 with some but only few CQ prescriptions thereafter, the mortality study was not performed in the CQ group due to lack of power.

### RA/SLE/SS study

The RA/SLE/SS study included subjects with and without rheumatoid arthritis (RA), systemic lupus erythematous (SLE), and Sjögren’s syndrome (SS), respectively. As in the Matched study on CQ and HCQ, we identified subjects with a diagnosis of RA, SLE, or SS, respectively, and matched them 1:1, 3:1, and 2:1 to controls, respectively, on sex and age (rounded to integer years), sampled without replacement.

### Medication and comorbidity

Using Anatomical Therapeutic Chemical (ATC) codes, we identified use of CQ and HCQ. SLE, RA, SS, and ischemic heart disease were defined using the International Classification of Diseases, 10^th^ revision (ICD-10). Diabetes mellitus, hypertension, and congestive heart failure were all defined using a combination of ICD-10 and ATC codes at baseline as done previously.(11) Exact definitions using ICD-10 and ATC codes are given in the **Supplementary Material**. Use of QT-prolonging drugs was defined as concomitant prescription of one of the 63 drugs listed on CredibleMeds.org on April 30^th^ 2020 as having a known risk of Torsade des Pointes (TdP, a possibly lethal polymorphic form of ventricular tachycardia) except for CQ and HCQ (see the **Supplementary Material** for the full list).

### Statistical considerations

For subjects in the paired population, subjects served as their own controls and the results were thus inherently adjusted for comorbidity, age, and sex. We adjusted for a change in use of QT-prolonging drugs where appropriate. We used a random-effects regression model for continuous variables.

For subjects in the matched population, we compared continuous electrocardiographic variables using linear regression crudely as well as with adjustment for RA, SLE, SS, use of QT-prolonging drugs, hypertension, ischemic heart disease congestive heart failure, and diabetes mellitus, as appropriate. Categorical variables were tested using a two-sample test of proportions.

For subjects in the mortality population, we used Cox regression with full adjustment as with the matched population, as appropriate. Subjects were censored upon emigration individually or 300 days after the last claimed prescription for each case/control pair. We used time-on-study as underlying time variable in the Cox model.

The Danish Data Protection Agency approved the use of de-identified data (2007-58-0015) on the conditions that the exact number of subjects in groups of < 3 subjects not be disclosed and that no calculations (including p-values) be reported on groups of < 5 subjects. All estimates were reported along with a 95 % confidence interval indicated as (lower to upper). Data management and statistical computations were performed in Stata (version 16), and a p-value < 0.05 was considered significant.

## Results

### Paired study

We identified ten subjects who had an ECG taken during a treatment of CQ and who had a prior ECG without treatment of CQ (**Table 1**, CQ population). Similarly, for treatment with HCQ, we identified 32 subjects (**Table 1**, HCQ population).

**Table 1.** Paired population: ECG findings in subjects with ECG during a treatment with chloroquine or hydroxychloroquine. Reported are findings from a prior baseline ECG and change upon drug administration.

| Variables | ECG before and during treatment |  |
| --- | --- | --- |
|  | Chloroquine | Hydroxychloroquine |
| n | 10 | 32 |
| Age, years | 61.6 ± 5.6 | 54.5 ± 12.1 |
| Sex, Females | <3 | 24 (75 %) |
| Baseline QT-prolonging drug aside from CQ/HCQ | <3 | 4 (13 %) |
| Study QT-prolonging drug aside from CQ/HCQ | <3 | 6 (19 %) |
| **ECG variables** |  |  |
| Baseline Heart Rate, bpm | 64.8 ± 14.7 | 70.9 ± 14.8 |
| ΔHeart Rate, bpm | 1.3 [-5.6;8.2], p=0.7 | 2.3 [-1.7;6.2], p=0.3 |
| Baseline QTcB, ms | 427 ± 30 | 432 ± 32 |
| ΔQTcB, ms | 6.0 [-0.6;12.7], p=0.08 | 2.4 [-6.3;11], p=0.6 |
| ΔQTcB > 30 ms | <3 | 3 (9 %) |
| Baseline QTcFrh, ms | 421 ± 20 | 420 ± 23 |
| ΔQTcFrh, ms | 5.5 [0.7;10], p=0.03 | 1.0 [-5.6;7.5], p=0.8 |
| ΔQTcFrh > 30 ms | <3 | <3 |
| Baseline JTc, ms | 316 ± 22 | 317 ± 22 |
| ΔJTc, ms | 2.8 [-3.3;8.9], p=0.4 | -0.6 [-6.3;5.0], p=0.8 |
| Baseline QRS duration, ms | 104 ± 21 | 98 ± 18 |
| ΔQRS duration, ms | 2.2 [-1.4;5.8], p=0.2 | 0.2 [-3.7;4.0], p=0.9 |
| Baseline PR interval, ms | 164 ± 24 | 164 ± 29 |
| ΔPR interval, ms | -0.4 [-9.1;8.2],<br>p=0.9 | -2.8 [-9.4;3.8],<br>p=0.4 |
| Baseline RBBB | <3 | <3 |
| Study RBBB | <3 | <3 |
| Baseline LBBB | <3 | <3 |
| Study LBBB | <3 | <3 |

Compared to their baseline ECG, subjects taking CQ had a significantly longer QTcFrh by 5.5 ms (0.7 to 10, p=0.03). We found no evidence of an increased QTcFrh after use of HCQ (1.0 ms, −5.6 to 7.5 ms, p=0.8).

### Matched study

We identified 28 and 172 subjects with an ECG taken during a period of treatment with CQ or HCQ, respectively, and matched them 10:1 with controls.

Subjects on CQ (**Table 2**), compared to controls, were more likely to suffer from ischemic heart disease, but not hypertension. With full adjustment for RA, SLE, SS, QT-prolonging drugs, hypertension, diabetes, ischemic heart disease, and congestive heart failure, QTcFrh was 4.7 ms longer in the CQ group compared to control, but with a wide confidence interval (−3.4 to 13 ms, p=0.3).

**Table 2.**
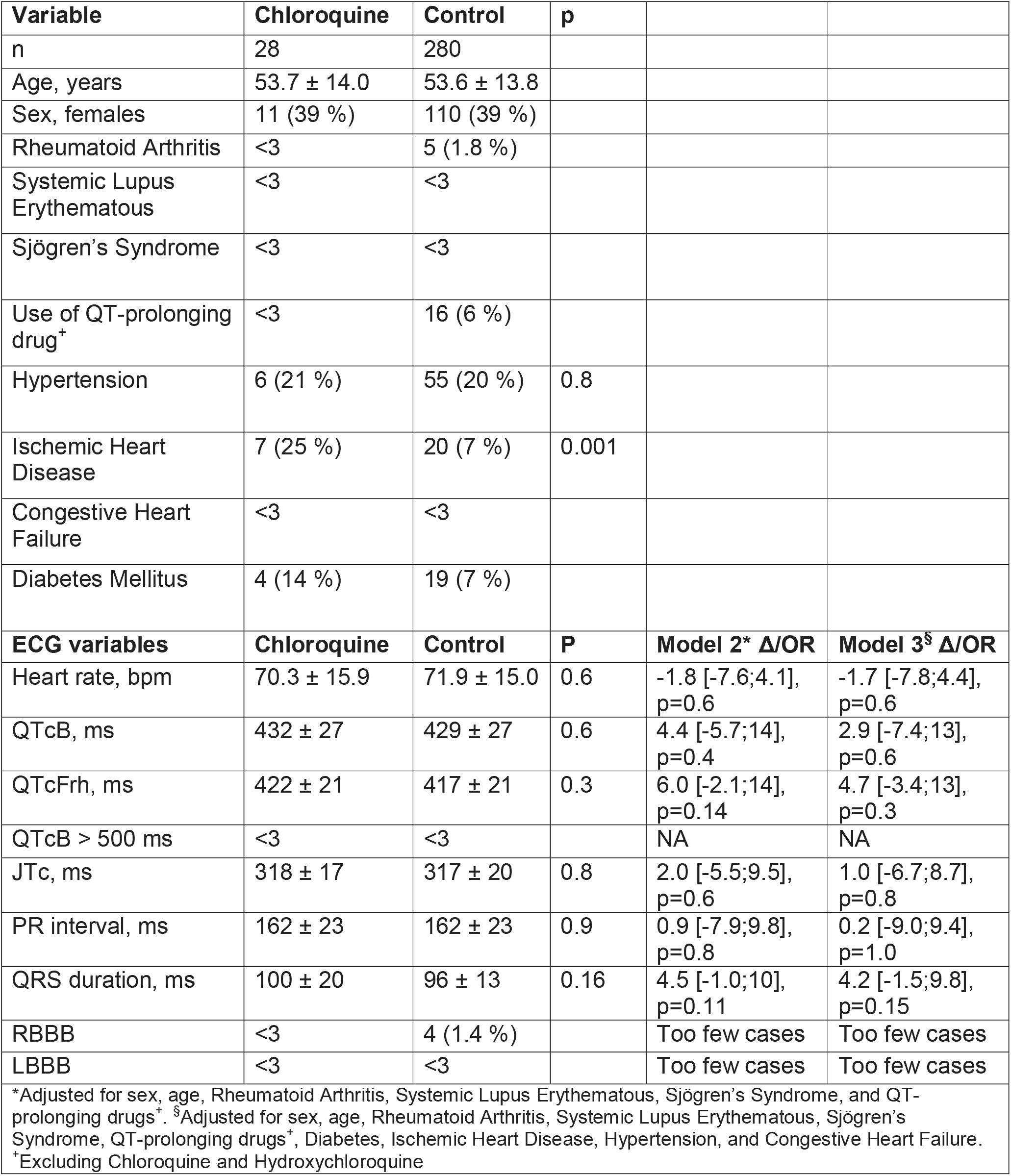
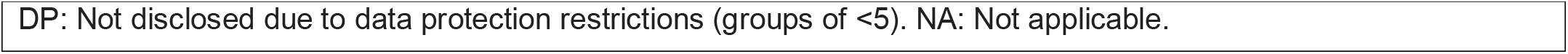
Matched population: Characteristics and ECG findings in subjects taking chloroquine vs. control. For the ECG findings, adjusted models show effects independent of comorbidities.

| Variable | Chloroquine | Control | p |  |  |
| --- | --- | --- | --- | --- | --- |
| n | 28 | 280 |  |  |  |
| Age, years | 53.7 ± 14.0 | 53.6 ± 13.8 |  |  |  |
| Sex, females | 11 (39 %) | 110 (39 %) |  |  |  |
| Rheumatoid Arthritis | <3 | 5 (1.8 %) |  |  |  |
| Systemic Lupus Erythematosus | <3 | <3 |  |  |  |
| Sjögren's Syndrome | <3 | <3 |  |  |  |
| Use of QT-prolonging drug<sup>+</sup> | <3 | 16 (6 %) |  |  |  |
| Hypertension | 6 (21 %) | 55 (20 %) | 0.8 |  |  |
| Ischemic Heart Disease | 7 (25 %) | 20 (7 %) | 0.001 |  |  |
| Congestive Heart Failure | <3 | <3 |  |  |  |
| Diabetes Mellitus | 4 (14 %) | 19 (7 %) |  |  |  |
| ECG variables | Chloroquine | Control | P | Model 2* Δ/OR | Model 3<sup>§</sup> Δ/OR |
| Heart rate, bpm | 70.3 ± 15.9 | 71.9 ± 15.0 | 0.6 | -1.8 [-7.6;4.1],<br>p=0.6 | -1.7 [-7.8;4.4],<br>p=0.6 |
| QTcB, ms | 432 ± 27 | 429 ± 27 | 0.6 | 4.4 [-5.7;14],<br>p=0.4 | 2.9 [-7.4;13],<br>p=0.6 |
| QTcFrh, ms | 422 ± 21 | 417 ± 21 | 0.3 | 6.0 [-2.1;14],<br>p=0.14 | 4.7 [-3.4;13],<br>p=0.3 |
| QTcB > 500 ms | <3 | <3 |  | NA | NA |
| JTc, ms | 318 ± 17 | 317 ± 20 | 0.8 | 2.0 [-5.5;9.5],<br>p=0.6 | 1.0 [-6.7;8.7],<br>p=0.8 |
| PR interval, ms | 162 ± 23 | 162 ± 23 | 0.9 | 0.9 [-7.9;9.8],<br>p=0.8 | 0.2 [-9.0;9.4],<br>p=1.0 |
| QRS duration, ms | 100 ± 20 | 96 ± 13 | 0.16 | 4.5 [-1.0;10],<br>p=0.11 | 4.2 [-1.5;9.8],<br>p=0.15 |
| RBBB | <3 | 4 (1.4 %) |  | Too few cases | Too few cases |
| LBBB | <3 | <3 |  | Too few cases | Too few cases |

\*Adjusted for sex, age, Rheumatoid Arthritis, Systemic Lupus Erythematosus, Sjögren's Syndrome, and QT-prolonging drugs<sup>+</sup>. <sup>§</sup>Adjusted for sex, age, Rheumatoid Arthritis, Systemic Lupus Erythematosus, Sjögren's Syndrome, QT-prolonging drugs<sup>+</sup>, Diabetes, Ischemic Heart Disease, Hypertension, and Congestive Heart Failure. <sup>+</sup>Excluding Chloroquine and Hydroxychloroquine
DP: Not disclosed due to data protection restrictions (groups of <5). NA: Not applicable.

Subjects on HCQ (**Table 3**) more often than controls had RA, SLE or SS, as well as hypertension, ischemic heart disease, congestive heart failure, and diabetes. In the unadjusted analysis, we found an increased QTcFrh interval of 5 ms (p=0.002). However, after adjustment for RA, SLE, SS and QT-prolonging drugs, there was no significant difference in QTcFrh between cases and controls (3.1 ms, −1.1 to 7.3 ms, p=0.15) and after additional adjustment for ischemic heart disease, hypertension, diabetes, and congestive heart failure, the difference in QTcFrh was reduced further to 1.3 ms (−2.9 to 5.6, p=0.5). QRS duration followed a similar pattern in the adjusted and unadjusted analyses, whereas the JT interval also in the unadjusted analysis was comparable between the groups.

**Table 3.**
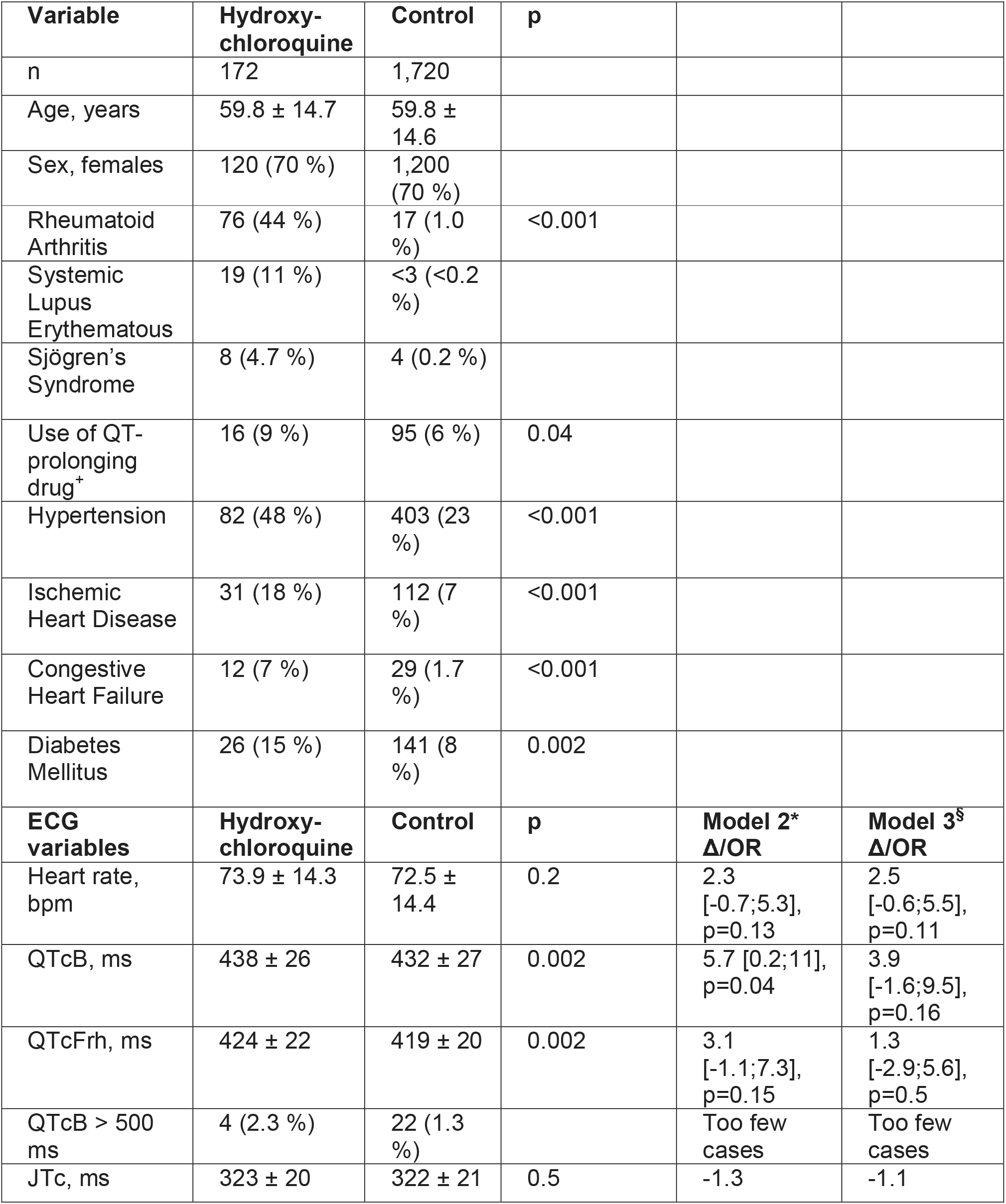
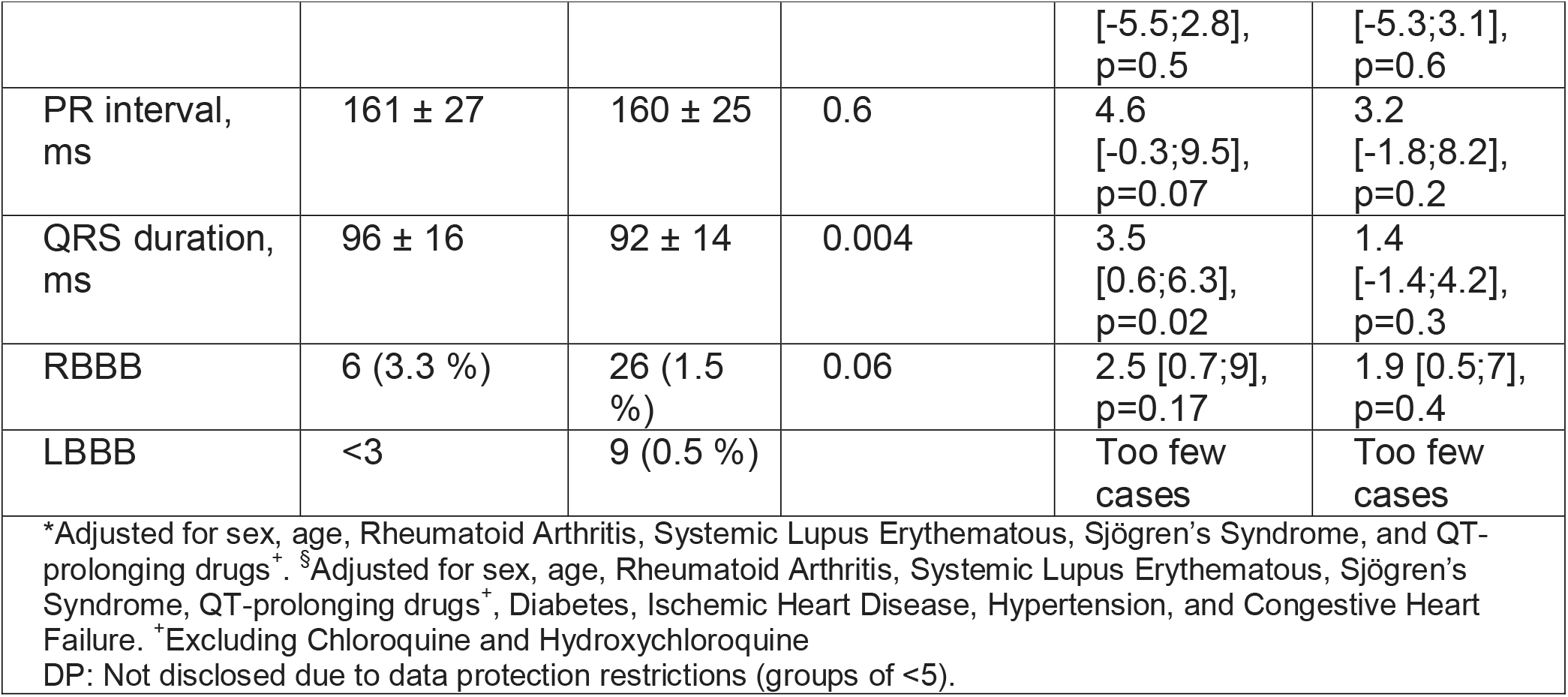
Matched population: Characteristics and ECG findings in subjects taking hydroxychloroquine vs. control. For the ECG findings, adjusted models show effects independent of comorbidities.

### Mortality study

We identified 3,368 subjects with a prescription for HCQ irrespective of an ECG (**Table 4**).

**Table 4.**
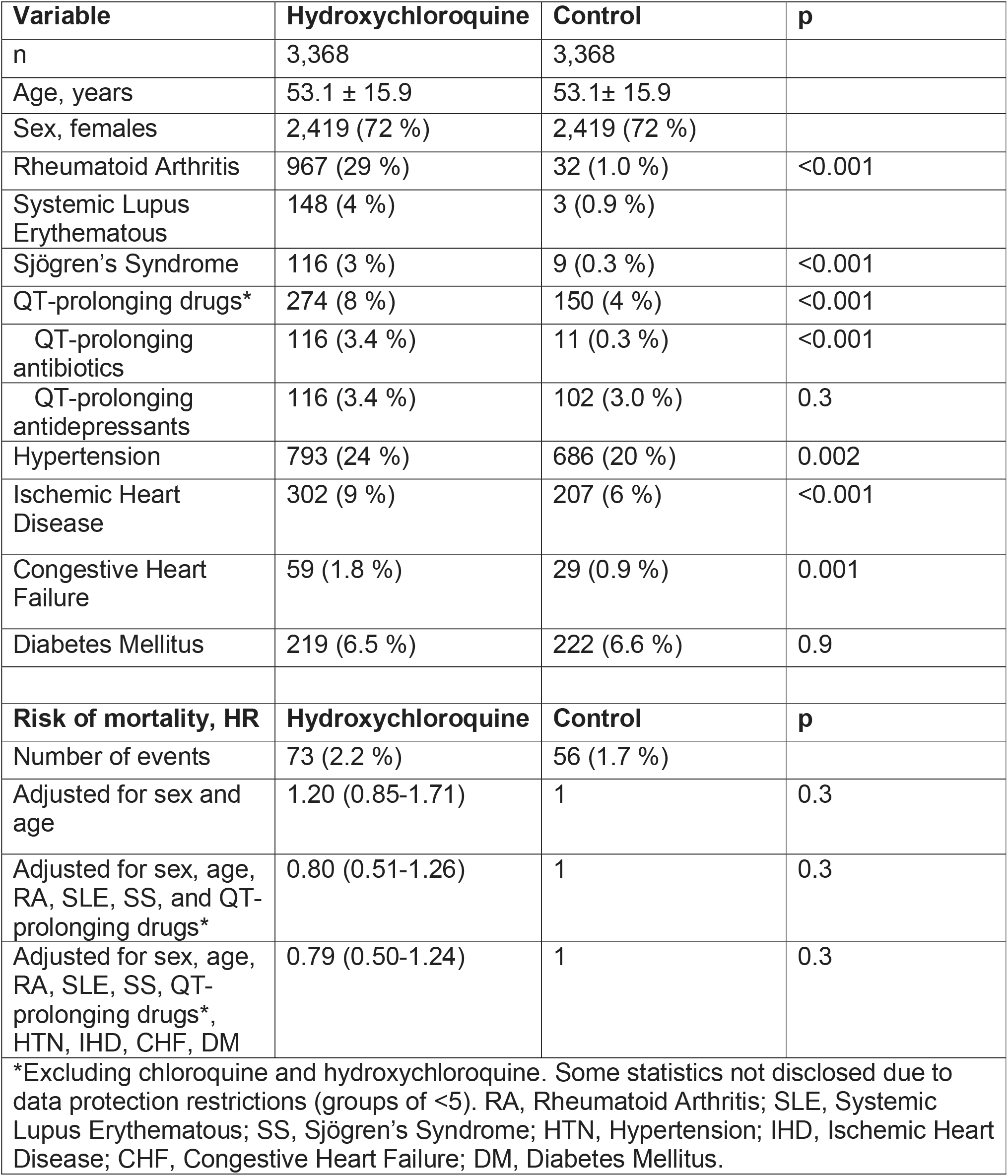
Mortality population: Baseline characteristics and mortality in subjects taking hydroxychloroquine compared to sex- and age-matched controls.

Subjects on HCQ were more likely to have RA, SLE, SS, hypertension, ischemic heart disease, and congestive heart failure. In the most crude analysis, adjusted only for sex and age, mortality was not different between subjects on HCQ and controls (HR=1.20, 0.85 to 1.71, p=0.3). With adjustment for RA, SLE, SS, and use of other QT-prolonging drugs, we still found no significant difference between Hydroxychloroquine and matched controls (HR=0.80, 0.51 to 1.26, p=0.3). Further adjustment for ischemic heart disease, diabetes, hypertension, and congestive heart failure did not change the association materially. Kaplan-Meier survival curves are shown in **Figure 1**.

**Figure 1.**
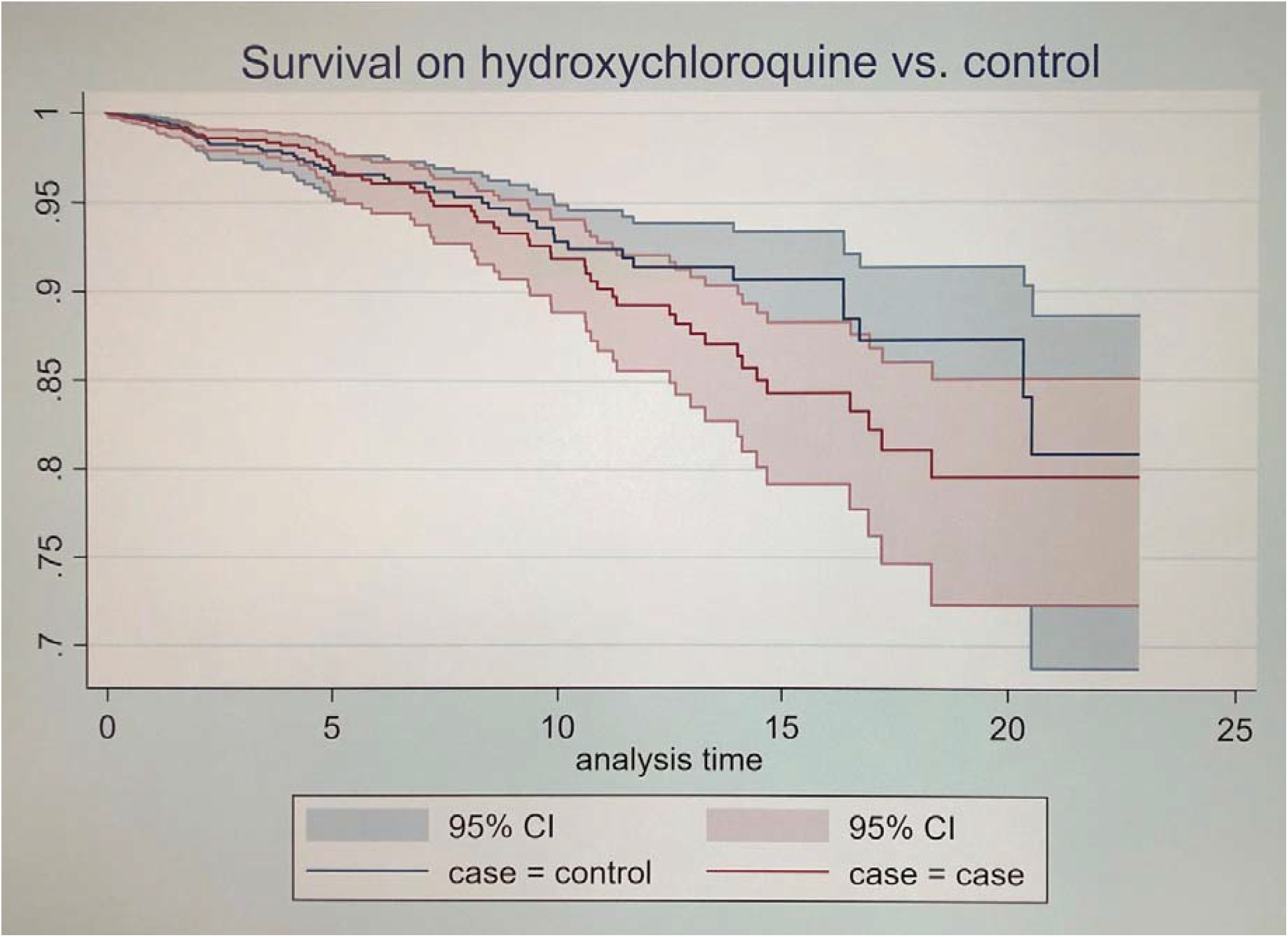
Survival for people on hydroxychloroquine and matched controls.

### RA/SLE/SS study

In the RA/SLE/SS study we defined three populations of cases and controls with and without RA, SLE, and SS respectively. We found no difference in crude or adjusted QTcFrh compared to control in any of the populations (**Supplementary Tables 1-3**). Heart rate was significantly higher in subjects with RA and SLE, but lower in subjects with SS. Crude PR interval was increased in subjects with SS, but not with adjustment for diabetes, ischemic heart disease, hypertension, and congestive heart failure. In the fully adjusted adjusted for sex, age, SS, SLE, QT-prolonging drugs, ischemic heart disease, diabetes, hypertension, and congestive heart failure, patients with RA had a shorter QRS duration compared to controls.

## Discussion

In this real-world study, we found a small, but significant, heart rate-corrected QT-prolongation of 5 ms with CQ, but no QT prolongation with HCQ. We did not find increased mortality in subjects taking CQ or HCQ.

### Electrocardiographic safety

CQ and HCQ are both listed as known causes of TdP on CredibleMeds.org,(12) and CQ is recognized by the World Health Organization (WHO) as prolonging the QT/QTc interval.(5) However, WHO states that CQ is associated with a low risk of cardiotoxicity based on PK/PD modelling.(5) In the present study, we found an increase in QTcFrh of 5.5 ms in the paired CQ population, and no difference the paired HCQ population. Another study(10) showed that a 1,500 mg dose of CQ yielded an increase in QTc of 15-30 ms.

Matched on sex and age, subjects receiving HCQ showed a 5 ms increase in QTcFrh. However, the increased QTc disappeared upon adjustment for use of QT-prolonging medication and connective tissue diseases. Hence, the increased QTcFrh was likely caused by rheumatic disease and not HCQ. Thus, in both the matched and the paired analyses, we were unable to demonstrate any statistically significant increase in QTc associated with use of HCQ.

### Mortality

We did not find that use of HCQ was associated with an increased mortality. This is not surprising, since the toxicology profile of HCQ is better than that of CQ.(13) WHO reports that the few deaths associated with use of HCQ were caused by overdosing and chronic indications for HCQ.(5)

Only three case studies have demonstrated arrhythmic adverse events in association with use of CQ/HCQ. One study(7) involved a syncope in a patient with SLE and end stage renal disease and QT prolongation before HCQ initiation. Another study(8) documents TdP in a patient with SLE, with a history of cirrhosis, HBV-related hepatoma, prior myocardial infarction, and ventricular septal defect. The third study(9) involved a patient with SLE, who also suffered from Congestive Heart Failure, Chronic Kidney Disease stage 5, and Hypertension. All three case studies thus features patients with severe QT-prolonging comorbidity and chronic rather than episodic use of Hydroxychloroquine. Collectively, evidence from the present and other studies found little risk of death or arrhythmic events associated with episodic use of HCQ.

### Indication for HCQ

Malaria was and still remains an indication for HCQ in Denmark, although it is no longer recommended.(14) HCQ is commonly prescribed for connective tissue diseases such as Rheumatoid Arthritis, Systemic Lupus Erythematous, and Sjögren’s Syndrome. We were only able to identify the reason for the HCQ treatment in forty to sixty percent of the cases, likely because milder connective tissue cases were treated in primary care and therefore never obtained a hospital ICD-10 diagnosis detectable in our registries. Furthermore, some subjects may have been prescribed HCQ as malaria prophylaxis.

### CQ and HCQ for treatment of COVID-19

CQ/HCQ has emerged as a possible treatment for COVID-19.(15) Naturally, given the known risk of increased QTc, concerns about the safety of the drugs have been raised.(16, 17) We have found different electrocardiographic safety profiles for CQ and HCQ. With CQ, we have demonstrated an increased QTcFrh of 5.5 ms. With HCQ, we found no increase in QTcFrh. For use of CQ and HCQ, respectively, we found no increased mortality.

CQ and HCQ have been used as malaria prophylaxis for decades without raising red flags and the use of HCQ in higher doses to treat auto-immune diseases also has not been associated with an increased mortality. The few case studies that exist were associated with chronic use of HCQ beyond the duration corresponding to a COVID-19 treatment and with severe QT-prolonging comorbidity.

Arrhythmias are commonly seen in patients with COVID-19 (one study(18) reports 17 %) independently of treatment with CQ, HCQ, or azithromycin. The virus likely attacks the myocardium directly due to the high prevalence of angiotensin converting enzyme 2 (ACE2) receptors in the heart.(19) Shi et al. found cardiac lesions (based on high-sensitivity Troponin I) in 20 % of COVID-19 patient on admission.(20) There is no solid evidence for any significant proarrhythmic effects of CQ or HCQ. However, COVID-19 may play a super additive role in the safety profile of CQ/HCQ, and that can only be assessed with randomized studies on CQ/HCQ.

The present study found no grounds for concern for HCQ and minor cautions for CQ in patients free of COVID-19. Saleh et al.(21) in 200 COVID-19 patients treated with CQ/HCQ plus 60 % azithromycin, found no cases of TdP, although seven patients discontinued treatment due to QT-prolongation. Mercuro et al.(16) found one case of TdP in 90 patients treated with HCQ or HCQ+azithromycin. Bessière et al.(17) found no cases of TdP in 40 patients treated with HCQ plus/minus azithromycin. A limitation of the present study is that we cannot exclude an interaction between COVID-19 and CQ/HCQ treatment, and thus further studies – preferably randomized clinical trials – are needed to assess the cardiac safety of CQ/HCQ in patients with COVID-19.

## Conclusions

Using matched and paired studies of subjects free of COVID-19 receiving chloroquine or hydroxychloroquine, we found that use of chloroquine but not hydroxychloroquine was associated with a small increase in QTc. We were unable to show increased mortality associated with hydroxychloroquine.

## Data Availability

Data is not publically available due to patient confidentiality and data protection.

## Supplemental material for

### Supplemental methods

Use of Chloroquine was defined when a prescription for Chloroquine (ATC code: P01BA01) was claimed.

Use of Hydroxycloroquine was defined in the same way using ATC code P01BA02.

Systemic Lupus Erythematous was defined as any of the ICD-10 codes DM32, DG058A, DG737C, DI328B, DI398C, DJ991C, or DN085A, or either ICD-8 code 695.4 or 734.1.

Rheumatoid Arthritis was defined as any of the ICD-10 codes DM05 or DM06, or any of the ICD-8 codes 712.0, 712.1, or 712.3.

Ischemic heart disease was defined as any of the ICD-10 codes I20, I21, I23, I24, I25 or the ICD-8 code 410.

Diabetes mellitus was defined as any of the ICD-10 codes E10, E11, E12, E13 or E14, or use of insulin (ATC: A10A) or oral antidiabetics (ATC: A10B), or the ICD-8 code 761.1.

Hypertension was present if any of the ICD-10 codes I10 or I15, or the ICD-8 code 401 was found, or if we found concurrent use of two of these classes of medication: alpha blockers (ATC: C02A, C02B, C02C), non-loop diuretics (ATC: C02L, C03A, C03B, C03D, C03E, C03X, C07X, C07C, C07D, C08G, C02DA, C09BA, C09DA, C09XA52), vasodilators (ATC: C02DB, C02DD, C02DG, C04, C05), beta blockers (ATC: C07), calcium blockers (ATC: C08, C09BB, C09DB), or angiotensin converting enzyme inhibitors (ATC: C09).

Heart failure was present if the ICD-10 codes I42, I50, or J81, or the ICD-8 code 427.0 was found, or if a prescription of loop diuretics (ATC: C03C) was found.

The following drugs and ATC codes were considered QT-prolonging in the present study. The list was taken from CredibleMeds.org on April 30, 2020 (revision March 19, 2020), and is comprised of those drugs categorized as having a known risk of Torsade de Pointes (TdP), with the omissions of Chloroquine and Hydroxychloroquine.

| **Generic drug name** | **ATC code(s)** |
| --- | --- |
| Aclarubicin | L01DB04 |
| Amiodarone | C01BD01 |
| Anagrelide | L01XX35 |
| Arsenic trioxide | L01XX27 |
| Astemizole | R06AX11 |
| Azithromycin | J01FA10, S01AA26, J01RA07 |
| Bepridil | C08EA02 |
| Cesium Chloride | No ATC available |
| Chlorpromazine | N05AA01 |
| Chlorprothixene | N05AF03 |
| Cilostazol | B01AC23 |
| Ciprofloxacin | J01MA02, S01AE03, S02AA15, S03AA07, J01RA10, J01RA11, J01RA12 |
| Cisapride | A03FA02 |
| Citalopram | N06AB04 |
| Clarithromycin | J01FA09, A02BD04, A02BD05, A02BD06, A02BD07, A02BD09, A02BD11 |
| Cocaine | N01BC01, R02AD03, S01HA01, S02DA02 |
| Disopyramide | C01BA03 |
| Dofetilide | C01BD04 |
| Domperidone | A03FA03 |
| Donepezil | N06DA02, N06DA52, N06DA53 |
| Dronedarone | C01BD07 |
| Droperidol | N05AD08 |
| Erythromycin | D10AF02, J01FA01, S01AA17, D10AF52 |
| Escitalopram | N06AB10 |
| Flecainide | C01BC04 |
| Fluconazole | D01AC15, J02AC01 |
| Gatifloxacin | J01MA16, S01AE06 |
| Grepafloxacin | J01MA11 |
| Halofantrine | P01BX01 |
| Haloperidol | N05AD01 |
| Hydroquinidine | C01BA13 |
| Ibogaine | No ATC available |
| Ibutilide | C01DB05 |
| Levofloxacin | A02BD10, J01MA12, S01AE05, J01RA05 |
| Levomepromazine | N05AA02 |
| Levomethadyl acetate | N07BC03 |
| Levosulpride | N05AL07 |
| Mesoridazine | N05AC03 |
| Methadone | N07BC02, N02AC52 |
| Moxifloxacin | J01MA14, S01AE07 |
| Nifekalant | Not marketed in Denmark |
| Ondansetron | A04AA01 |
| Oxaliplatin | L01XA03 |
| Papaverine HCl | A03AD01, G04BE02, G04BE52 |
| Pentamidine | P01CX01 |
| Pimozide | N05AG02 |
| Probucol | C10AX02 |
| Procainamide | C01BA02 |
| Propofol | N01AX10 |
| Quinidine | C01BA01, C01BA51, C01BA71 |
| Roxithromycin | J01FA06 |
| Sevoflurane | N01AB08 |
| Sotalol | C07AA07, C07FX02, C07BA07 |
| Sparfloxacin | J01MA09 |
| Sulpride | N05AL01 |
| Sultopride | N05AL02 |
| Terfenadine | R06AX12 |
| Terlipressin | H01BA04 |
| Terodiline | G04BD05 |
| Thioridazine | N05AC02 |
| Vandetanib | L01XE12 |

## Supplementary Tables

**Supplementary Table 1.** Characteristics and ECG findings in people with Rheumatoid Arthritis.

| Variable | Rheumatoid Arthritis | Control | p |  |  |
| --- | --- | --- | --- | --- | --- |
| n | 6,329 | 6,329 |  |  |  |
| Age, years | 64.2 ± 15.6 | 64.2 ± 15.6 |  |  |  |
| Sex, females | 4,766 (75%) | 4,766 (75%) |  |  |  |
| Systemic Lupus Erythematosus | 102 (1.6 %) | 7 (0.1 %) | <0.001 |  |  |
| Sjögren's Syndrome | 144 (2.3 %) | 22 (0.4 %) | <0.001 |  |  |
| Use of QT-prolonging drugs<sup>#</sup> | 619 (9.8 %) | 414 (6.5 %) | <0.001 |  |  |
| Hypertension | 2,436 (38 %) | 1,816 (29 %) | <0.001 |  |  |
| Ischemic Heart Disease | 902 (14 %) | 531 (9 %) | <0.001 |  |  |
| Congestive Heart Failure | 370 (5.9 %) | 186 (2.7 %) | <0.001 |  |  |
| Diabetes Mellitus | 775 (12 %) | 443 (7 %) | <0.001 |  |  |
| ECG variables | Rheumatoid Arthritis | Control | p | Model 2*<br>Δ/OR | Model 3<sup>s</sup><br>Δ/OR |
| Heart rate, bpm | 73.6 ± 14.4 | 73.0 ± 14.2 | 0.01 | 0.6 [0.1;1.1,<br>p=0.01 | 0.6 [0.1;1.1],<br>p=0.02 |
| QTcB, ms | 434 ± 27 | 433 ± 27 | 0.07 | 0.6 [-0.3;1.5],<br>p=0.2 | -0.2 [-1.1;0.8],<br>p=0.7 |
| QTcFrh, ms | 420 ± 22 | 420 ± 22 | 0.4 | 0.1 [-0.7;0.9],<br>p=0.8 | -0.6 [-1.4;0.2],<br>p=0.13 |
| QTcB > 500 ms | 97 (1.5 %) | 96 (1.5 %) | 0.9 | 1.0 [0.7;1.3],<br>p=0.9 | 0.8 [0.6;1.1],<br>p=0.2 |
| JTc, ms | 322 ± 23 | 322 ± 22 | 0.7 | -0.1 [-0.8;0.7],<br>p=0.8 | -0.1 [-0.9;0.6],<br>p=0.7 |
| PR interval, ms | 162 ± 27 | 161 ± 27 | 0.4 | 0.4 [-0.5;1.3],<br>p=0.4 | -0.4 [-1.3;0.5],<br>p=0.4 |
| QRS duration, ms | 93 ± 16 | 93 ± 16 | 0.2 | -0.1 [-0.6;0.4],<br>p=0.7 | -0.8 [-1.2;-0.2],<br>p=0.01 |
| RBBB | 141 (2.2 %) | 133 (2.1 %) | 0.6 | 1.1 [0.8;1.4],<br>p=0.6 | 1.0<br>[0.8;1.3],<br>p=0.9 |
| LBBB | 45 (0.7 %) | 60 (1.0 %) | 0.14 | 0.8 [0.5;1.1],<br>p=0.19 | 0.7<br>[0.5;1.1],<br>p=0.09 |
| <sup>#</sup>Except Chloroquine and Hydroxychloroquine. <sup>*</sup>Adjusted for sex, age, Sjögren's Syndrome, Systemic Lupus Erythematosus, and QT-prolonging drugs <sup>#</sup>. <sup>§</sup>Adjusted for sex, age, Sjögren's Syndrome, Systemic Lupus Erythematosus, Diabetes, Ischemic Heart Disease, Hypertension, and Congestive Heart Failure. |  |  |  |  |  |

**Supplementary Table 2.** Characteristics and ECG findings in people with Systemic Lupus Erythematous

| Variable | Systemic Lupus Erythematosus | Control | p |  |  |
| --- | --- | --- | --- | --- | --- |
| n | 540 | 1,620 |  |  |  |
| Age, years | 56.4 ± 15.0 | 56.4 ± 15.0 |  |  |  |
| Sex, females | 465 (86%) | 1,395 (86 %) |  |  |  |
| Rheumatoid Arthritis | 97 (18 %) | 18 (1.1 %) | <0.001 |  |  |
| Sjögren's Syndrome | <3 | 9 (0.6 %) |  |  |  |
| Use of QT-prolonging drugs<sup>#</sup> | 54 (10 %) | 96 (5.9 %) | 0.001 |  |  |
| Hypertension | 192 (36 %) | 355 (22 %) | <0.001 |  |  |
| Ischemic Heart Disease | 71 (13 %) | 109 (7 %) | <0.001 |  |  |
| Congestive Heart Failure | 23 (4.3 %) | 29 (1.8 %) | 0.001 |  |  |
| Diabetes Mellitus | 40 (7.4 %) | 85 (5.3 %) | 0.06 |  |  |
| ECG variables | Systemic Lupus Erythematosus | Control | p | Model 2*<br>Δ/OR | Model 3<sup>s</sup><br>Δ/OR |
| Heart rate, bpm | 73.4 ± 14.1 | 71.8 ± 13.0 | 0.02 | 1.3 [-0.1;2.6],<br>p=0.06 | 1.3 [-0.1;2.6],<br>p=0.07 |
| QTcB, ms | 433 ± 24 | 431 ± 25 | 0.08 | 2.0 [-0.5;4.5],<br>p=0.11 | 1.1 [-1.4;3.6],<br>p=0.4 |
| QTcFrh, ms | 420 ± 19 | 419 ± 20 | 0.5 | 0.7 [-1.3;2.7],<br>p=0.5 | -0.1 [-2.1;1.9],<br>p=0.9 |
| QTcB > 500 ms | 3 (0.6 %) | 11 (0.7 %) |  | Too few cases | Too few cases |
| JTc, ms | 324 ± 20 | 324 ± 21 | 0.9 | 0.4 [-1.6;2.4],<br>p=0.7 | 0.1 [-2.0;2.2],<br>p=0.9 |
| PR interval, ms | 158 ± 24 | 157 ± 25 | 0.6 | 1.2 [-1.4;3.7],<br>p=0.4 | 0.4 [-2.2;3.0],<br>p=0.8 |
| QRS duration, ms | 90 ± 13 | 90 ± 12 | 1.0 | -0.2 [-1.4;1.1],<br>p=0.8 | -0.7 [-1.9;0.5],<br>p=0.3 |
| RBBB | 4 (0.7 %) | 14 (0.9 %) |  | Too few cases | Too few cases |
| LBBB | <3 | 9 (0.6 %) |  | Too few cases | Too few cases |
| <sup>#</sup>Except Chloroquine and Hydroxychloroquine. <sup>*</sup>Adjusted for sex, age, Rheumatoid Arthritis, Sjögren's Syndrome, and QT-prolonging drugs<sup>#</sup>. <sup>§</sup>Adjusted for sex, age, Rheumatoid Arthritis, Sjögren's Syndrome, QT-prolonging drugs<sup>#</sup>, Diabetes, Ischemic Heart Disease, Hypertension, and Congestive Heart Failure.<br>DP: Not disclosed due to data protection restrictions (groups of <5). |  |  |  |  |  |

**Supplementary Table 3.** Characteristics and ECG findings in people with Sjögren’s Syndrome

| Variable | Sjögren's Syndrome | Control | p |  |  |
| --- | --- | --- | --- | --- | --- |
| n | 1,174 | 2,348 |  |  |  |
| Age, years | 64.3 ± 14.1 | 64.3 ± 14.1 |  |  |  |
| Sex, females | 1,042 (89%) | 2,084 (89 %) |  |  |  |
| Rheumatoid Arthritis | 168 (14 %) | 41 (1.8 %) | <0.001 |  |  |
| Systemic Lupus Erythematosus | 54 (4.6 %) | 6 (0.3 %) | <0.001 |  |  |
| Use of QT-prolonging drugs<sup>#</sup> | 132 (11 %) | 151 (6.4 %) | <0.001 |  |  |
| Hypertension | 467 (40 %) | 677 (29 %) | <0.001 |  |  |
| Ischemic Heart Disease | 184 (16 %) | 198 (8.4 %) | <0.001 |  |  |
| Congestive Heart Failure | 46 (3.9 %) | 67 (2.9 %) | 0.09 |  |  |
| Diabetes Mellitus | 116 (9.9%) | 170 (7.2 %) | 0.007 |  |  |
| ECG variables | Sjögren's Syndrome | Control | p | Model 2*<br>Δ/OR | Model 3<sup>§</sup><br>Δ/OR |
| Heart rate, bpm | 72.3 ± 13.4 | 73.3 ± 13.7 | 0.04 | -1.3<br>[-2.3;-0.3],<br>p=0.009 | -1.2<br>[-2.2;-0.2],<br>p=0.01 |
| QTcB, ms | 433 ± 26 | 433 ± 26 | 0.4 | -1.1 [-3.0;0.7],<br>p=0.2 | -1.6<br>[-3.4;0.3],<br>p=0.10 |
| QTcFrh, ms | 421 ± 21 | 420 ± 21 | 0.6 | 0.2 [-1.3;1.7],<br>p=0.8 | -0.3<br>[-1.8;1.3],<br>p=0.7 |
| QTcB > 500 ms | 8 (0.7 %) | 30 (1.3 %) | 0.11 | 0.6 [0.3;1.2],<br>p=0.16 | 0.5<br>[0.2;1.1],<br>p=0.09 |
| JTc, ms | 325 ± 22 | 323 ± 21 | 0.16 | 0.9 [-0.7;2.4],<br>p=0.3 | 0.7<br>[-0.8;2.3],<br>p=0.4 |
| PR interval, ms | 162 ± 25 | 160 ± 26 | 0.02 | 2.0 [0.2;3.9],<br>p=0.03 | 1.2<br>[-0.6;3.1],<br>p=0.20 |
| QRS duration, ms | 91 ± 15 | 91 ± 14 | 0.5 | -0.1 [-1.2;0.9],<br>p=0.8 | -0.5<br>[-1.5;0.5],<br>p=0.3 |
| RBBB | 24 (2.0 %) | 37 (1.6 %) | 0.3 | 1.3 [0.8;2.2],<br>p=0.3 | 1.2<br>[0.7;2.1],<br>p=0.5 |
| LBBB | 9 (0.8 %) | 25 (1.1 %) | 0.4 | 0.7 [0.3;1.6],<br>p=0.5 | 0.7<br>[0.3;1.6],<br>p=0.4 |
| <sup>#</sup>Except Chloroquine and Hydroxychloroquine. <sup>*</sup>Adjusted for sex, age, Rheumatoid Arthritis, Systemic Lupus Erythematosus and QT-prolonging drugs<sup>#</sup>. <sup>§</sup>Adjusted for sex, age, Rheumatoid Arthritis, Systemic Lupus Erythematosus, QT-prolonging drugs<sup>#</sup>, Diabetes, Ischemic Heart Disease, Hypertension, and Congestive Heart Failure. |  |  |  |  |  |

